# Testing Claimed Associations of Prenatal Acetaminophen with Neurodevelopmental Disorders: Re-analysis Using Bias-Correction Methods

**DOI:** 10.64898/2026.08.28.26361666

**Authors:** Ambika Grover, Joana Reis-Pardal, Randall J. Ellis, John P.A. Ioannidis, Chirag J. Patel

## Abstract

**Importance:** A 2025 systematic review concluded there was "strong evidence of a likely relationship" between prenatal acetaminophen exposure and neurodevelopmental disorders (NDDs), directly informing a federal health advisory, based on qualitative synthesis without quantitative pooling or bias correction.

**Objective:** To determine, via meta-analysis, whether the association claimed between prenatal acetaminophen exposure and attention-deficit/hyperactivity disorder (ADHD), autism spectrum disorder (ASD), and other NDDs survives publication-bias correction and sensitivity methods applied to the same underlying studies.

**Data Sources:** The 46 studies identified in a 2025 Navigation Guide systematic review, based on a PubMed search (through February 25, 2025) supplemented by Web of Science and Google Scholar searches that yielded no additional studies; no additional search was performed for this reanalysis.

**Study Selection:** Of the 46 studies, 24 (52%) reported a ratio measure (hazard ratio, odds ratio, relative risk, or incidence rate ratio) and were retained; studies reporting only continuous outcomes were excluded.

**Data Extraction and Synthesis:** Estimates were collapsed to 1 per study, outcome group, and design to correct a unit-of-analysis error in the original review. Random-effects meta-analysis was used. We assessed eight bias-correction methods, a credibility ceiling analysis, and 5 quality-filtered subsets, then repeated the analyses after harmonizing odds and hazard ratios onto a common risk ratio scale.

**Main Outcomes and Measures:** Pooled ratio and 95% CI for ADHD, ASD, and other NDDs under each method, before and after harmonization.

**Results:** Among 30 estimates from 24 studies (sample sizes, 307-2,480,797), the ASD association lost statistical significance (pooled ratio, 1.08; 95% CI, 0.97-1.19) from corrected weighting alone, before any further bias correction. The ADHD association remained positive under most methods but lost statistical significance under a 15% credibility ceiling. The other NDD association was not robust under bias correction, reversing sign under 1 method and failing a test for evidential value (p-curve, P = .71).

**Conclusions and Relevance:** The claim of "strong evidence" for an acetaminophen-NDD association was not supported by meta-analysis of the same studies. A modest, method-dependent ADHD signal persisted with most methods, but the ASD and other-NDD associations did not survive bias-correction and sensitivity methods.

**Key Points:** 

**Question:** Does the "strong evidence" verdict linking prenatal acetaminophen exposure to attention-deficit/hyperactivity disorder (ADHD) and autism spectrum disorder (ASD) survive standard meta-analytic and bias-correction methods applied to the same underlying studies?

**Findings:** In this meta-analysis of 30 study-level estimates from a 2025 systematic review,^1^ the ASD association was not significant after corrected study weighting alone, and the association with other neurodevelopmental disorders was not robust under scrutiny. The ADHD association survived most, but not all, of eight publication-bias-correction methods and was not statistically significant under a modest 15% credibility ceiling.

**Meaning:** These findings suggest that the original review’s claim of strong evidence linking prenatal acetaminophen use to neurodevelopmental disorders is not supported by quantitative reanalysis of the same underlying studies.

---

Systematic reviews face a methodological choice: to pool underlying studies quantitatively, accept the assumptions that pooling requires, or synthesize them qualitatively, accepting the subjectivity that qualitative synthesis requires.^2^ A recent review of prenatal acetaminophen neurodevelopmental disorders (NDDs) chose the latter path, applying the Navigation Guide framework (a review methodology developed for environmental-health evidence) to 46 studies spanning attention-deficit/hyperactivity disorder (ADHD) (20 studies), autism spectrum disorder (ASD) (8 studies), and other NDDs (18 studies).^1,3^ The review avoided meta-analysis, citing substantial heterogeneity in exposure assessment, outcome definition, and confounder adjustment across studies. Instead, it constructed a qualitative synthesis using a numeric scoring system the authors devised, including a risk-of-bias rubric and a separate strength-of-evidence rubric criterion. That review concluded that there was “strong evidence” of a likely relationship between prenatal acetaminophen exposure and both ADHD and ASD.

The strong causal verdict raised considerable debate. The review informed a federal health advisory.^4^ Many professional bodies reasserted their stance not to consider acetaminophen as being harmful in pregnancy.^5,6^ An umbrella review of the 9 systematic reviews on this topic rated 7 as critically low and 2 (including Prada et al.) as low confidence by AMSTAR 2.^7^

Publication and selective-reporting bias are major threats that may easily invalidate observed signals of epidemiological associations.^8,9^ Familial confounding is another major threat for this specific topic. Among the studies included in the systematic review, two analyses re-estimated the association within families, accounting for shared genetics and shared family environment.^10,11^ Both showed the association attenuating toward the null. Sibling and co-twin comparison designs strengthen epidemiologic inference by implicitly controlling for genetic and environmental factors shared within families, effectively reducing familial confounding.^12,13^

Here, we reanalyze the contested systematic review, using the same underlying studies, weighting each study once rather than by its number of sub-analyses, and applying eight publication-bias-correction methods and a credibility-ceiling sensitivity analysis. We also examine five "higher-quality" subsets built from the risk-of-bias and strength-of-evidence scores the original review already assigned to every study. Finally, we repeat all analyses after converting odds ratios and hazard ratios onto a common risk-ratio scale, since the pooled studies report hazard ratios, odds ratios, relative risks, and incidence rate ratios that are only approximately equivalent. We ask whether the review’s "strong evidence" verdict survives this scrutiny (Figure 1).

**Figure 1:**
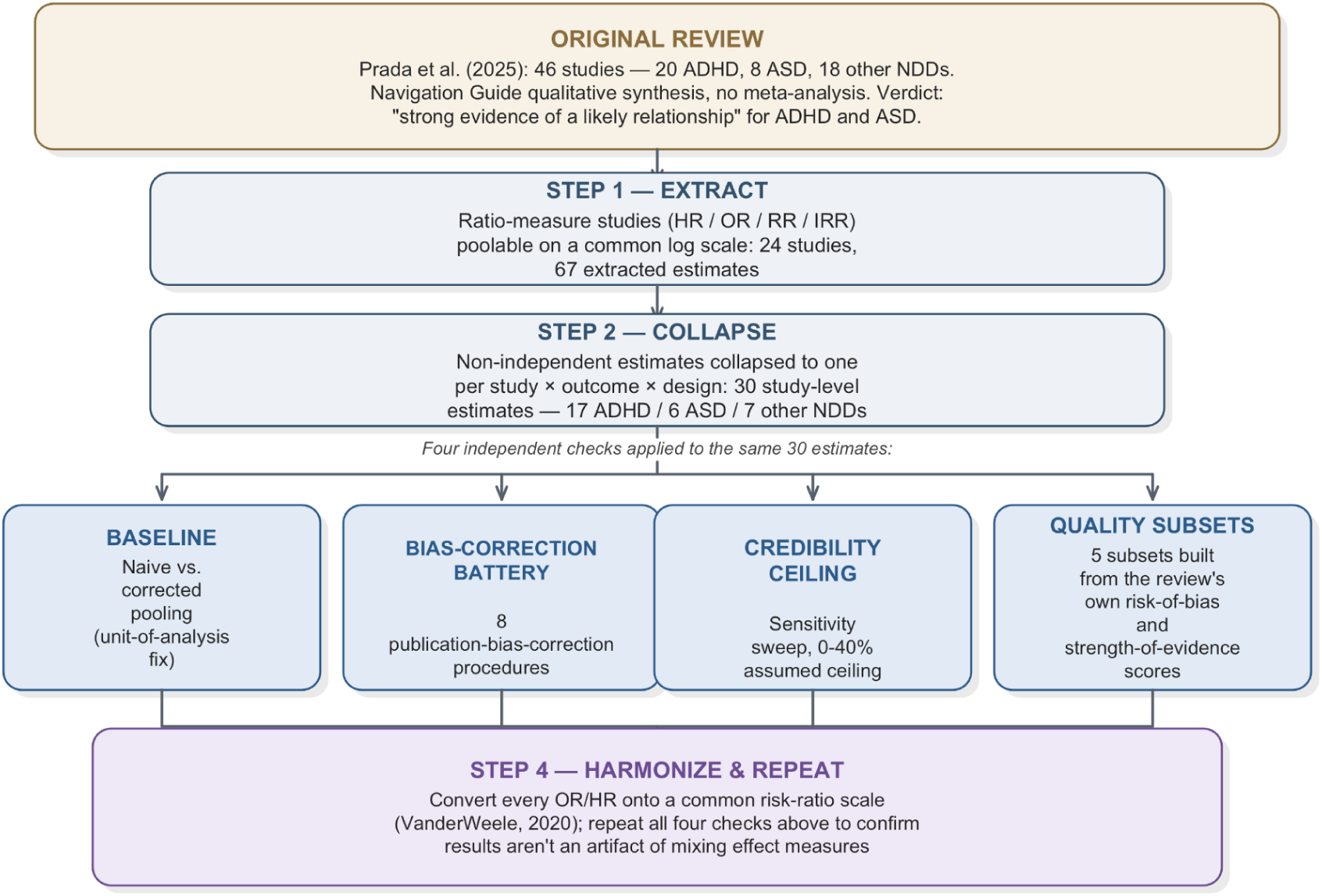
Schematic of the full reanalysis. The original review’s 46 studies are narrowed to 30 study-level estimates (17 attention-deficit/hyperactivity disorder [ADHD], 6 autism spectrum disorder [ASD], 7 other neurodevelopmental disorders [NDDs]), then stress-tested in four independent ways, twice: once on the data as reported, once after converting all ratio measures onto a common scale.

## METHODS

### Study identification and data extraction

We extracted every study reported in the original review’s outcome tables (46 studies total) that reports a ratio measure (hazard ratio, odds ratio, relative risk, incidence rate ratio), since these sit on a common log scale and can be pooled. Studies reporting continuous outcome measures (regression coefficients, mean differences, IQ point differences) were excluded, as they constitute a separate outcome family that cannot be pooled with ratio measures without additional standardization steps that may require tenuous assumptions. Cross-verification against the original review’s own tables separately surfaced one apparent error in the source material itself; see eMethods 1.

The 24 retained studies varied in how exposure was ascertained: most relied on maternal self-report or interview,^11,14–30^ while a smaller number used objective measures: fetal or maternal biomarkers^31–33^, prescription/dispensing records,^34^ or a mixed exposure definition.^10,35^

### Handling non-independent estimates within a study

Several studies report multiple estimates from the same underlying cohort— different outcome subscales, different exposure-duration windows, different assessment timepoints. Treating each as an independent data point would inflate that study’s influence on any pooled estimate in proportion to how many sub-analyses it reports. This may manufacture false precision by combining correlated estimates as though they were independent replications. To address this, we collapsed each study to one estimate per study, per outcome group, per analytic design, using the unweighted mean of the log-transformed estimates and the largest (most conservative) standard error among that study’s own contributing rows, rather than a combination that would implicitly assume independence. As a sensitivity check, we repeated this collapse using the smallest (least conservative) standard error instead; pooled point estimates were materially unchanged for all three outcome groups, with only modest narrowing of confidence intervals and shifts in p-values (eTable 2). Where a study reported both a population-average estimate and a sibling-controlled estimate, we retained these as two separate rows rather than averaging them, since they estimate different quantities; namely, a population-average effect and a within-family effect. These are not interchangeable sub-analyses of the same estimand.^10,11^ This yielded 30 study-level estimates: 17 for ADHD, 6 for ASD, and 7 for other NDDs.

### Publication-bias-correction methods

We compiled candidate publication-bias-correction methods from two published reviews of such methodology, then restricted to widely cited (>100 Scopus citations) methods with actively maintained software implementations, yielding eight methods (eMethods 2).^36–43^

The eight procedures applied make different assumptions.^40–48^ Precision-based methods include the trim-and-fill,^39^ which detects funnel-plot asymmetry, imputes the effect sizes of studies that asymmetry implies are missing, and recomputes the pooled estimate including them; and limit meta-analysis,^44^ which models the relationship between study precision and effect size and extrapolates to the estimate implied at infinite precision. Selection modeling methods include the Vevea-Hedges weight-function selection model,^41–43^ which uses pre-specified p-value intervals with different selection weights; the Dear & Begg selection model^45^ where the cut-points and shape of the selection function are computed from the data; p-uniform,^47^ which derives an effect-size estimate using only the subset of statistically significant studies; and the Mathur-VanderWeele method,^48^ which reports corrected estimates under assumed publication-selection ratios (here we used ratio=2 and 4) in favor of “affirmative” studies, and, separately, the minimum such ratio (the "S-value") required to reduce the pooled estimate to non-significance. The Copas selection model^46^ assumes publication is driven by an underlying selection score correlated with the study’s effect, and the chance of publication is explicitly modeled as a function of the study’s precision; we report results at 70% and 40% publication rates (representing modest and severe selection) among fits with goodness-of-fit P > .10, and the publication rate where statistical significance is lost. Finally, the p-curve^40^ can bias-correct continuous measures, but here, for ratio measures, we use it as an evidential value check (whether the distribution of significant p-values supports genuine underlying effects) rather than a bias-corrected estimate.

### Credibility-ceiling sensitivity analysis

The credibility ceiling method^49^ considers that no single non-randomized study, however large, can be more than (1−c) certain about the direction of an underlying effect, given the potential for unmeasured confounding inherent to observational designs. For each ceiling c, we inflated a study’s variance only enough to bring its implied certainty down to (1-c), unless already below (1-c). We varied c from 0% to 40%.

### Quality-filtered subsets

For every included study, the original report presents a risk-of-bias score and a strength-of-evidence score (eMethods 3).^1^ We transcribed these scores directly from the original review’s tables and constructed five subsets using criteria the review itself already reports: risk-of-bias score of 1, the review’s own lowest risk rating; “strong” or “very strong” strength-of-evidence score; prospective cohort designs only, excluding case-control and nested case-control studies; sample size greater than 500; and exposure measured via biomarker or prescription registry.

### Effect-measure harmonization

Throughout the above, hazard ratios, odds ratios, relative risks, and incidence rate ratios were pooled on a common natural-log scale without further adjustment, defensible when the outcome is rare (conventionally under ∼15% prevalence at end of follow-up); both ADHD (∼8–10%) and ASD (∼2–3%) fall under that threshold.^50,51^ Because we cannot rule out that this approximation affects some pooled estimates, we repeated all analyses converting every odds ratio and hazard ratio onto a directly comparable risk-ratio scale using an approximate conversion;^52^ relative risks and incidence rate ratios remained unconverted.

### Transparency

Figure 1 shows the analytical pipeline. This exploratory re-analysis and re-evaluation did not require ethical approval. Full data and code are available here.

## RESULTS

### Naive baseline and corrected weighting

A naive pooled analysis treating all 67 extracted rows as independent produced statistically significant positive associations for all three outcome groups: ADHD 1.31 (95% CI 1.22–1.41, p<10^-12^, from 15 studies contributing 34 rows), ASD 1.14 (95% CI 1.01–1.28, p=0.03, from 5 studies contributing 9 rows), and other NDDs 1.13 (95% CI 1.08-1.18, p<10^-8^, from 7 studies contributing 24 rows). This naive approach lets studies with more sub-analyses dominate the pooled estimate (eFigure 1): for ASD, one study’s three sub-analyses from a single 64,322-participant cohort carried 39.4% of total statistical weight, more than a separate 2.48-million-participant study’s two analytic arms combined (35.5%).

After collapsing each study into a single estimate per outcome group and analytic design for re-weighting, 30 studies remained (eTable 1). The ASD pooled estimate shrank and changed from nominally significant to non-significant, simply by correcting the study weighting.

Corrected-study level pooled estimates were for ADHD 1.33 (95% CI 1.18–1.49, p<10^-5^, 17 studies including 2 sibling-controlled arms), for ASD 1.08 (95% CI 0.97–1.19, p=0.17, 6 studies including 1 sibling-controlled arm), and for other NDDs 1.16 (95% CI 1.08–1.24, p<10^-4^, 7 studies, none sibling-controlled). Between-study heterogeneity (I²) was 92.7%, 81.8%, and 0.1%, respectively.

### Bias-correction methods (Table 1)

The ADHD association remained positive under every method, though the effect magnitude varied considerably, from 1.02 (weight-function selection model) to 1.31 (trim-and-fill). Statistical significance at p=0.05 was lost only for limit meta-analysis and the weight-function model; no assumed selection ratio reduced the Mathur-VanderWeele S-value estimate to non-significance, and p-curve showed evidential value.

**Table 1:** Pooled ratio and 95% confidence interval (CI) from each of the eight publication-bias-correction methods, applied to the study-level dataset as originally reported, both as originally reported and after effect-measure harmonization *ratio needed to yield non-significant association

| Method | ADHD (Unharmonized) | ADHD (Harmonized) | ASD (Unharmonized) | ASD (Harmonized) | Other NDDs (Unharmonized) | Other NDDs (Harmonized) |
| --- | --- | --- | --- | --- | --- | --- |
| 0. Naive (no correction) | 1.33<br>(1.18–1.49) | 1.20<br>(1.11–1.30) | 1.08<br>(0.97–1.19) | 1.05<br>(0.99–1.11) | 1.15<br>(1.08–1.24) | 1.11<br>(1.04–1.18) |
| 1. Trim-and-fill | 1.31<br>(1.17–1.47) | 1.09<br>(0.98–1.20) | 1.06<br>(0.95–1.18) | 1.04<br>(0.98–1.10) | 1.14<br>(1.07–1.22) | 1.09<br>(1.03–1.15) |
| **2. Limit meta-analyses** | 1.15<br>(0.99–1.33) | 1.11<br>(1.00–1.23) | 1.04<br>(0.93–1.17) | 1.04<br>(0.97–1.10) | 1.03<br>(0.90–1.18) | 1.05<br>(0.97–1.14) |
| **3. Weight-function selection model** | 1.02<br>(0.99–1.05) | 1.01<br>(1.00–1.03) | did not converge | did not converge | 1.00<br>(0.99–1.01) | did not converge |
| **4. Dear & Begg selection model** | 1.12<br>(1.03–1.36) | 1.13<br>(1.02–1.25) | 1.02<br>(0.99–1.14) | 1.02<br>(0.99–1.09) | 1.09<br>(1.01–1.19) | 1.06<br>(1.01–1.13) |
| **5. p-uniform** | 1.07<br>(1.06–1.08) | 1.05<br>(1.04–1.06) | 1.05<br>(1.00–1.09) | 1.03<br>(1.00–1.06) | 0.98<br>(0.65–1.19) | 1.00<br>(0.81–1.10) |
| **6. Mathur-VanderWeele (4x selection assumed)** | 1.25<br>(1.15–1.37) | 1.16<br>(1.08–1.23) | 1.04<br>(0.79–1.35) | 1.03<br>(0.84–1.25) | 1.13<br>(1.00–1.28) | 1.11<br>(1.01–1.23) |
| **Mathur-VanderWeele (2x selection assumed)** | 1.29<br>(1.18–1.41) | 1.18<br>(1.10–1.26) | 1.05<br>(0.88–1.27) | 1.04<br>(0.90–1.19) | 1.14<br>(1.03–1.27) | 1.11<br>(1.02–1.20) |
| **S-value\*** | not achievable at max. tested ratio | not achievable at max. tested ratio | not applicable, already non-significant | not applicable, already non-significant | 3.9 | not applicable, already non-significant |
| **7. Copas (70% published assumed)** | 1.25<br>(1.13–1.39) | 1.14<br>(1.06–1.22) | 1.05<br>(0.97–1.15) | 1.04<br>(0.99–1.08) | 1.14<br>(1.07–1.22) | 1.08<br>(1.04–1.13) |
| **Copas (40% published assumed)** | 1.17<br>(1.07–1.28) | 1.08<br>(1.03–1.13) | 1.04<br>(0.97–1.12) | 1.03<br>(0.99–1.08) | 1.11<br>(1.03–1.20) | 1.07<br>(1.02–1.12) |
| **Copas: publication rate at which significance is lost** | not lost in reliable range | not lost in reliable range | not applicable — already non-significant | not applicable — already non-significant | not lost in reliable range | not lost in reliable range |
| **8. p-curve (evidential-value test)** | right-skew test p=0.000 (evidential value present) | right-skew test p=0.000 (evidential value present) | right-skew test p=0.015 (evidential value present) | right-skew test p=0.015 (evidential value present) | right-skew test p=0.708 (no clear evidential value) | right-skew test p=0.708 (no clear evidential value) |

ASD remained non-significant under every method, with point estimates coming even closer to the null with all bias corrections (1.02-1.06) (Table 1). P-curve analysis gave a modest p=0.015.

For other NDDs, point estimates with bias-corrected methods ranged from 0.98 to 1.14 (the p-uniform method reversed the effect direction). Statistical significance was maintained for trim-and-fill, Dear & Begg, Copas (at both 70% and 40% assumed publication), and the Mathur-VanderWeele bound at η=2, but not for limit meta-analysis, the weight-function model, p-uniform, or the Mathur-VanderWeele bound at η=4. A p-curve analysis found no evidential value (p=0.71).

### Credibility ceiling sensitivity analysis (Figure 2)

ADHD lost statistical significance at the p<0.05 level at a ceiling of 15% and reached zero heterogeneity at c=16%; other NDDs lost statistical significance at 14% and reached near-zero heterogeneity at a markedly lower c=6%; ASD required no ceiling adjustment, since the estimate was already non-statistically significant.

**Figure 2:**
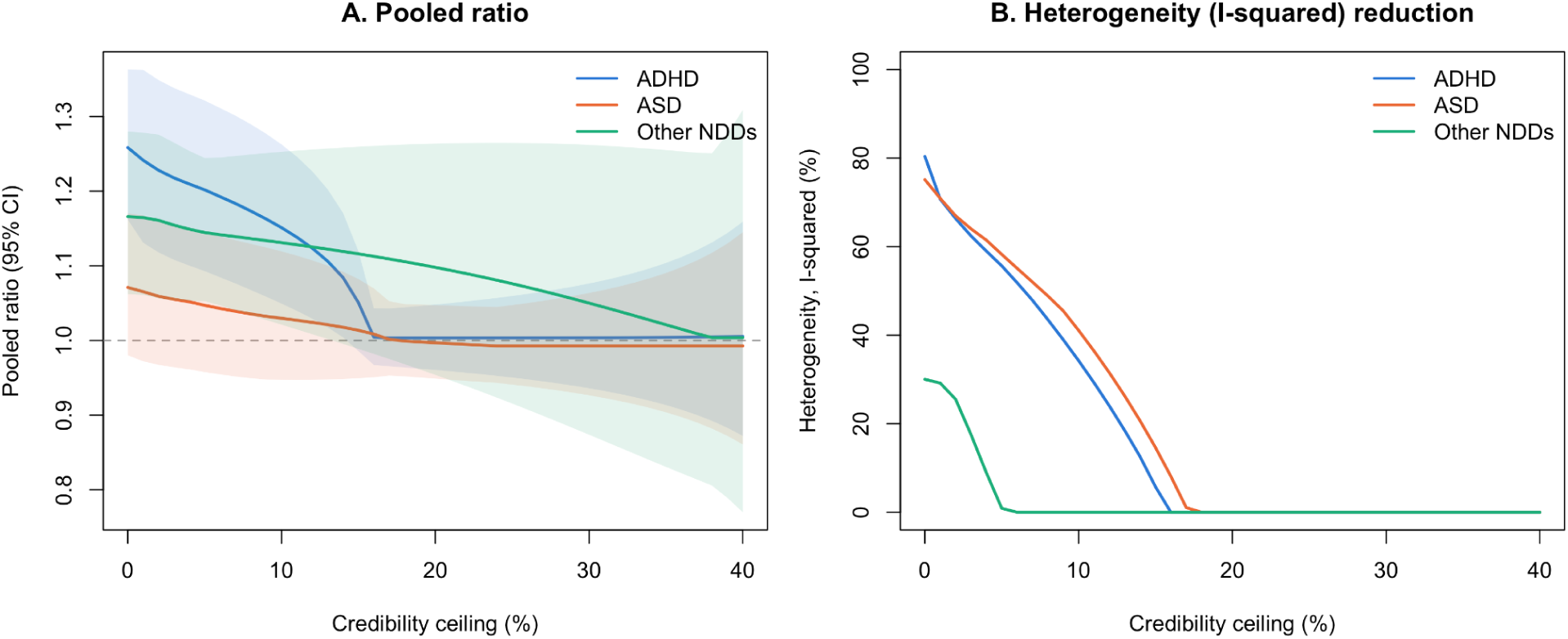
(A) Pooled ratio (with 95% confidence interval [CI] band) as a function of an assumed "credibility ceiling" — a cap on how much certainty any single non-randomized study is allowed to claim— swept from 0–40%, for each outcome group. ADHD crosses the null (ratio = 1) around a 16% ceiling; ASD requires no adjustment at all to already include the null; other NDDs cross at a 14% ceiling. (B) Between-study heterogeneity (I²) as a function of the same ceiling sweep. Other NDDs’ heterogeneity resolves at a markedly lower ceiling (∼6%) than ADHD’s and ASD’s (∼16–18%).

### Quality-filtered subsets (Figure 3)

For other NDDs, none of the five quality-filtered subsets meaningfully changed the pooled estimate relative to the full sample. For ADHD and ASD, however, filtering to the subsets the original review itself rated as lowest-risk-of-bias or highest-strength-of-evidence increased the pooled estimate relative to the full sample— for ADHD, from 1.33 (all studies) to 1.36 (low risk-of-bias subset) and 1.44 (strong strength-of-evidence subset); for ASD, from 1.08 (all studies, non-significant) to 1.21 (low risk-of-bias subset) and 1.15 (strong strength-of-evidence subset).

**Figure 3:**
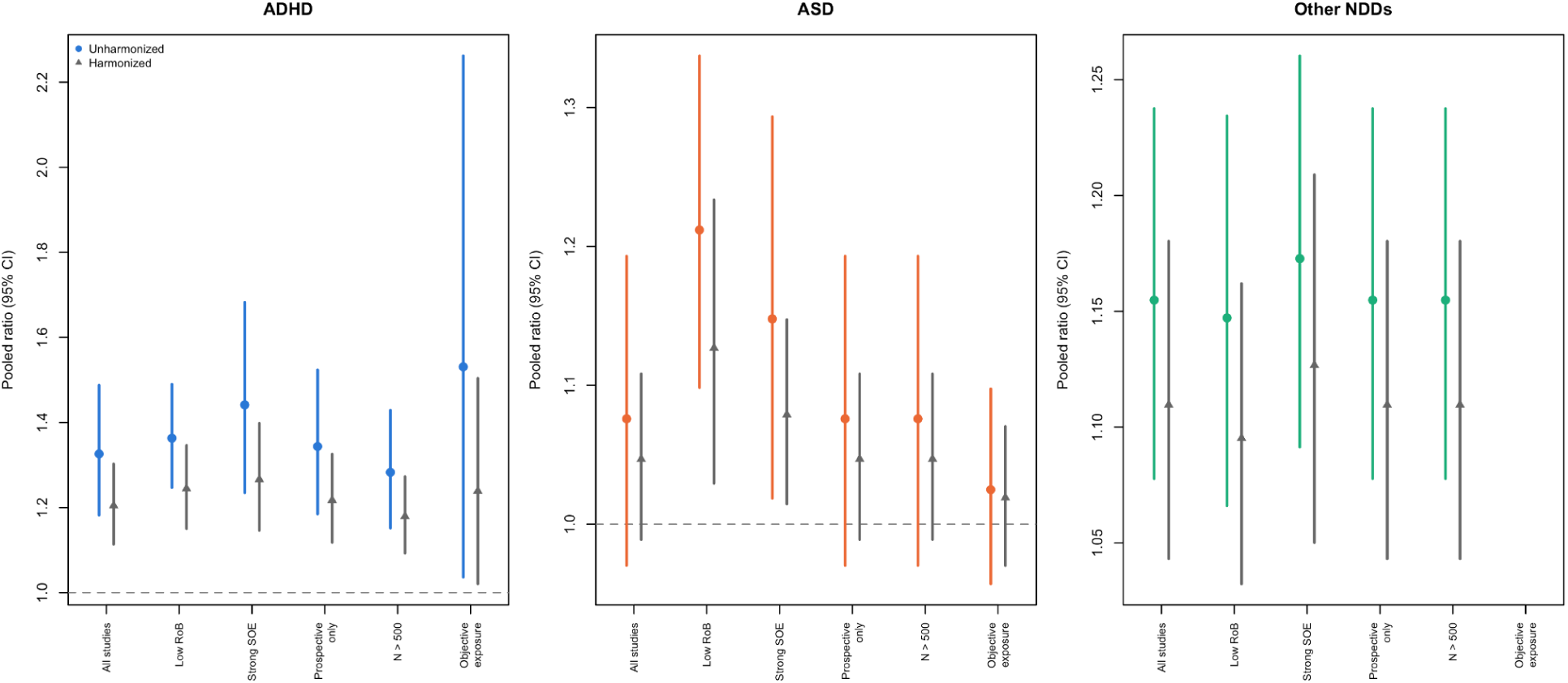
Pooled ratio across five subsets built entirely from the original review’s own risk-of-bias (RoB) and strength-of-evidence (SOE) scores, plus the full sample, unharmonized (circles) and harmonized (triangles). ADHD and ASD estimates climb rather than fall as the quality filter tightens, the opposite of what a working quality filter should produce, while other NDDs stay flat throughout. The ’Objective exposure’ set for Other NDDs reflects a single study (N=754) with an extremely wide confidence interval (not shown).

The mechanism for these changes is directly traceable in the original review’s own reported sub-scores. Both studies providing a sibling-controlled analysis also provide a population-average analysis of the same underlying cohort, scored separately by the original review. In both cases, the sibling-controlled arm scores lower than its own study’s population-average arm on the strength-of-evidence criterion for "large effect" and on "control of bias", while sibling-controlled designs represent the stronger approach for ruling out familial confounding. Filtering to the studies that the original review rated most highly therefore systematically removed the analyses best equipped to test the confounding explanation. This demonstrates circularity in the scoring instrument.

Notably, restricting the ASD analysis to the three studies measuring acetaminophen exposure objectively (via biomarker or prescription registry) rather than maternal self-report yielded a flat null estimate (1.02, 95% CI 0.96–1.10, p=0.48). The positive ASD association in this dataset is carried by studies relying on maternal recall of medication use, the exposure-assessment method that is most susceptible to differential recall bias when a mother’s child has received an autism diagnosis.

### Effect-measure harmonization

Measure-harmonized datasets produced consistently smaller effect sizes than their unharmonized counterparts. Naive pooled estimates of relative risks fell from 1.33 to 1.20 (ADHD), 1.08 to 1.05 (ASD), and 1.16 to 1.11 (other NDDs).

Bias-correction methods (Table 1) yielded largely similar results with the harmonized dataset as with the unharmonized counterpart for ASD and other NDDs, and made bias more visible for ADHD: trim-and-fill produced an estimate compatible with the null (1.09, 95% CI 0.98–1.20, versus 1.31 unharmonized) and imputed seven studies as missing from the funnel plot rather than one, revealing asymmetry that had been obscured while different effect measures were pooled uncorrected (Figure 4). Limit meta-analysis and weight-function selection models were also non-significant (eFigure 2).

**Figure 4:**
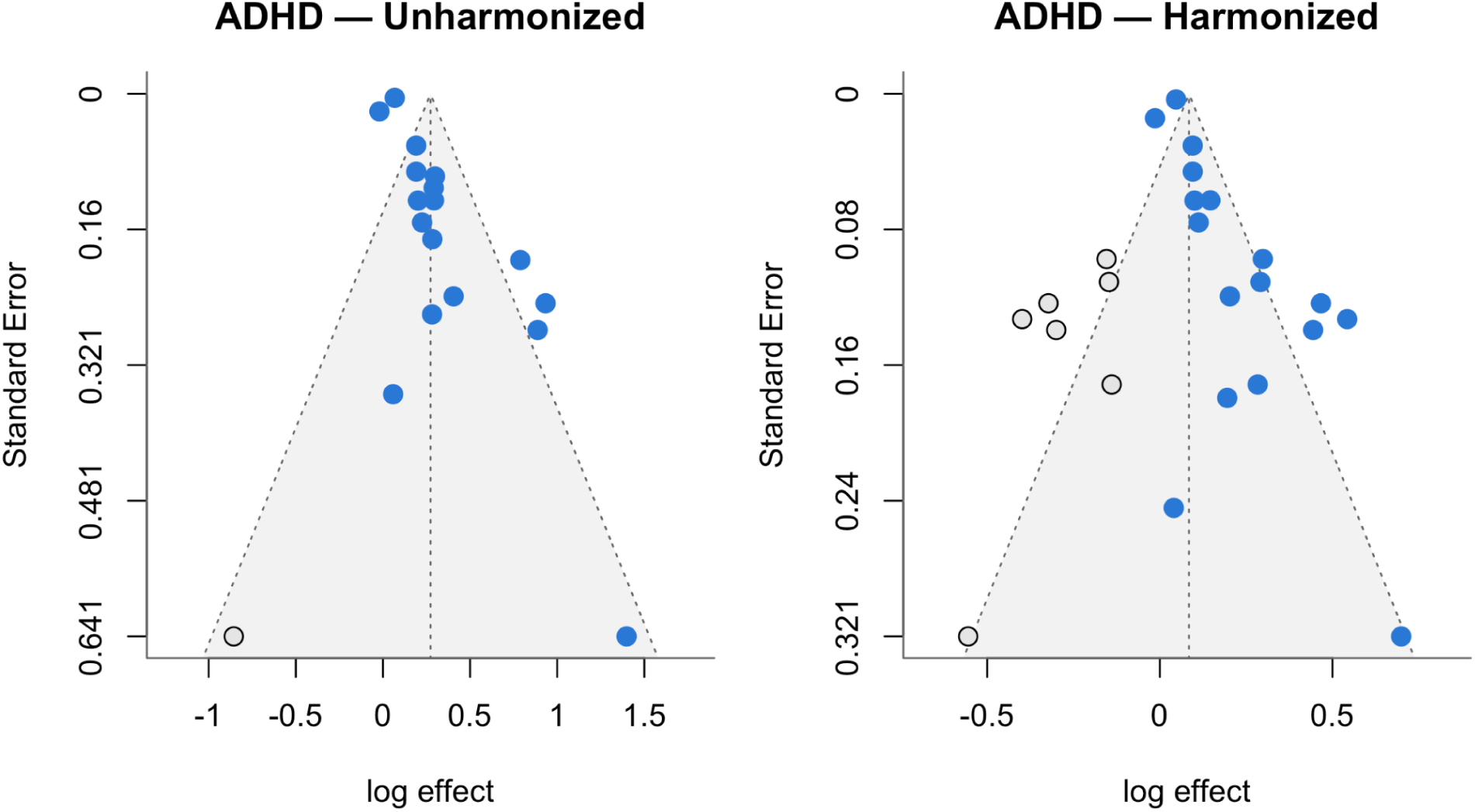
Funnel plot (each study’s effect size plotted against its precision) with trim-and-fill’s imputed "missing" studies shown as open circles, before (left) and after (right) converting odds ratios (OR) and hazard ratios (HR) onto a common risk-ratio scale. ADHD’s imputed-study count rises from 1 to 7 once measures are harmonized: an asymmetry (the classic signature of publication bias) that was present the whole time, just hidden while different effect measures were pooled together as if equivalent. The corresponding funnel plots for ASD and other NDDs appear in eFigure 3.

The credibility-ceiling thresholds were also essentially unchanged by harmonization: ADHD lost significance at a 15% ceiling both with and without harmonization, other NDDs at 14%, and ASD required no ceiling adjustment in either version.

The quality-subset scoring circularity replicated essentially unchanged (Figure 4): ADHD’s estimate still increased from 1.20 (all studies) to 1.24 (low risk-of-bias) to 1.27 (strong evidence), and ASD’s from 1.05 to 1.13 to 1.08 across the same three subsets. This appears to confirm that circularity is a property of the review’s scoring instrument, not an artifact of mixing effect measures.

## DISCUSSION

Reanalysis and reevaluation of evidence from studies of acetaminophen use and neurodevelopmental outcomes show that the observed associations are fragile. Association signals tend to disappear with better methodological choices that standardize and harmonize effect sizes across studies, and application of bias-correction methods further weakens claims of harmful effect. The ADHD association is the only one of the three outcomes that survives most corrections applied, but even then, its magnitude is method-dependent. Moreover, it loses statistical significance even with a modest credibility ceiling of 15%, i.e., assuming that no single observational study alone can offer more than 85% certainty for the presence of an association. The ASD association loses statistical significance even with the simple step of corrected study weighting alone, let alone any subsequent bias-correction method, and is entirely attributable to studies using a recall-dependent exposure measure. The association with other NDDs is the least robust, failing a direct test for evidential value and even reversing sign under at least one method.

Subsets of studies that were deemed to be of higher quality by the authors of the re-analyzed systematic review exhibit stronger signals for ADHD and ASD, but this is due to an identifiable bias. The original review’s own strength-of-evidence criteria treat larger effect sizes as evidence of higher study quality. This criterion systematically penalizes sibling-controlled analyses that are this literature’s strongest tool for ruling out familial confounding. A very large, recent sibling-matched analysis using data until 2026 also found adjusted hazard ratio of 1.00; 95% CI, 0.91-1.11 for ASD and 1.01; 95% CI, 0.93-1.08 for ADHD with consistently null associations across exposure timing, cumulative dose, and usage patterns, while positive associations were observed in conventional cohort analyses, and negative control analyses of pre-pregnancy exposure in the very same cohort.^53^ Familial residual confounding is a likely explanation for the persisting signal of association between acetaminophen use and neurodevelopmental outcomes. However, at least one relatively small sibling-controlled study^54^ reports potential neurodevelopmental harm, but it used continuous scale outcomes and thus was not included in the re-analysis.

This reanalysis should be read alongside prior meta-analyses of this literature. Masarwa et al. and Ricci et al. both report pooled estimates in a similar range (RR ≈1.34 for ADHD and 1.19 for ASD; OR 1.26 and 1.19, respectively) without publication-bias correction or measure-harmonization.^55,56^ Masarwa’s sensitivity analyses found the association highly susceptible to modest unmeasured confounding. A 2026 review by D’Antonio et al., meta-analyzing 17 of 43 included studies, found no association for any outcome among sibling-comparison studies, low-risk-of-bias studies, adjusted estimates, or studies with over five years of follow-up.^57^ Another recent review found a nominally significant ADHD association only under physician-based diagnosis (OR 1.17, 95% CI 1.08–1.27), but flagged high susceptibility to misclassification and found no signal for any other outcome.^58^

Some limitations should be discussed. First, we used data from the re-analyzed review and did not include more recent studies. Second, selective reporting on what types of adjusted or unadjusted analyses are published is impossible to identify in the absence of detailed protocol pre-registration, a practice that has not materialized in this field of observational epidemiology.^59^ Third, we did not attempt overall ranking of the evidence strength. However, one previous review using GRADE rated the evidence as low or very low.^58^ Another rating system for observational associations^58,60^ would rate the evidence as only suggestive. The factors that can decrease trustworthiness in observational studies of interventions are too numerous and contested.^61^

All the bias-correction methods that we employed have imperfect performance. The p-curve method can be susceptible to even limited bias in assessing observational, as opposed to interventional, evidence.^62^ Therefore, its detection of evidential value for ADHD and ASD may be spurious. Other bias-correction methods may also make assumptions that do not optimally capture the bias mechanisms in this field. Trim-and-fill, limit meta-analysis, and Copas selection methods assume that larger, rather than well-controlled, studies get preferentially published.

This is, however, exactly how population-level studies would be wrongly deemed more reliable than sibling-controlled ones. Therefore, the persistent signals for ADHD and other NDD with these methods may be spurious. Concurrently, precision-based methods may perform poorly and spuriously indicate bias when heterogeneity is substantial. Conversely, selection modeling methods can be sensitive to their specific assumptions, and the Vevea-Hedges model did not even converge in some analyses despite trying diverse options. Even the most parsimonious selection model (Mathur-VanderWeele) assumes that the published non-affirmative studies are representative of the unpublished ones, which may be a spurious assumption.

Allowing for these caveats, the specific claim of "strong evidence," is not supported by quantitative reanalysis of the same underlying studies to the degree the original synthesis concludes. Bias in the published literature on this topic may be substantial, and the methods we used may only partially detect and correct it. The authors of the re-analyzed review also note that alternatives to acetaminophen such as NSAID use in pregnancy or undertreated maternal fever carry their own risks. This is a genuine consideration on the other side of any precautionary calculus that should be weighed against, not independently of, the strength of the acetaminophen evidence itself.

## Article Information

### Data Sharing Statement

All code and data supporting this reanalysis are publicly available at https://github.com/ambika241/acetaminophen-ndd-reanalysis, including the full analytic pipeline, package-version lockfile (renv.lock), and every intermediate and final result reported in this manuscript.

### Additional Information

This study is an exploratory reanalysis of previously published data; no institutional review board approval was sought or required.

## Data Availability

All code and data supporting this reanalysis are publicly available at https://github.com/ambika241/acetaminophen-ndd-reanalysis

## References

1. Prada, D., Ritz, B., Bauer, A. Z. & Baccarelli, A. A. Evaluation of the evidence on acetaminophen use and neurodevelopmental disorders using the Navigation Guide methodology. Environ. Health 24, 56 (2025).

2. Ioannidis, J. P. A., Patsopoulos, N. A. & Rothstein, H. R. Reasons or excuses for avoiding meta-analysis in forest plots. BMJ 336, 1413–1415 (2008).

3. Woodruff, T. J. & Sutton, P. The Navigation Guide systematic review methodology: a rigorous and transparent method for translating environmental health science into better health outcomes. Environ. Health Perspect. 122, 1007–1014 (2014).

4. FACT: Evidence Suggests Link Between Acetaminophen, Autism. *The White House* https://www.whitehouse.gov/releases/2025/09/fact-evidence-suggests-link-between-acetaminophen-autism/ (2025).

5. Acetaminophen Use in Pregnancy and Neurodevelopmental Outcomes. https://www.acog.org/clinical/clinical-guidance/practice-advisory/articles/2025/09/acetaminophen-use-in-pregnancy-and-neurodevelopmental-outcomes.

6. Louwen, F. et al. Paracetamol (acetaminophen) use during pregnancy and autism risk: Evidence does not support causal association. Int. J. Gynaecol. Obstet. 171, 915–919 (2025).

7. Sheikh, J. et al. Maternal paracetamol (acetaminophen) use during pregnancy and risk of autism spectrum disorder and attention deficit/hyperactivity disorder in offspring: umbrella review of systematic reviews. BMJ 391, e088141 (2025).

8. Ioannidis, J. P. A. Why most published research findings are false. PLoS Med. 2, e124 (2005).

9. Viechtbauer, W. Publication bias in meta-analysis: Prevention, assessment and adjustments. Psychometrika 72, 269–271 (2007).

10. Ahlqvist, V. H. et al. Acetaminophen use during pregnancy and children’s risk of autism, ADHD, and intellectual disability. JAMA 331, 1205–1214 (2024).

11. Gustavson, K., et al. Acetaminophen use during pregnancy and offspring attention deficit hyperactivity disorder - a longitudinal sibling control study. JCPP Adv. 1, e12020 (2021).

12. McGue, M., Osler, M. & Christensen, K. Causal inference and observational research: The utility of twins: The utility of twins. Perspect. Psychol. Sci. 5, 546–556 (2010).

13. Lahey, B. B. & D’Onofrio, B. M. All in the family: Comparing siblings to test causal hypotheses regarding environmental influences on behavior. Curr. Dir. Psychol. Sci. 19, 319–323 (2010).

14. Alemany, S. et al. Prenatal and postnatal exposure to acetaminophen in relation to autism spectrum and attention-deficit and hyperactivity symptoms in childhood: Meta-analysis in six European population-based cohorts. Eur. J. Epidemiol. 36, 993–1004 (2021).

15. Leppert, B. et al. Association of maternal neurodevelopmental risk alleles with early-life exposures. JAMA Psychiatry 76, 834–842 (2019).

16. Liew, Z., Ritz, B., Rebordosa, C., Lee, P.-C. & Olsen, J. Acetaminophen use during pregnancy, behavioral problems, and hyperkinetic disorders. JAMA Pediatr. 168, 313–320 (2014).

17. Liew, Z., Ritz, B., Virk, J. & Olsen, J. Maternal use of acetaminophen during pregnancy and risk of autism spectrum disorders in childhood: A Danish national birth cohort study: Acetaminophen and Autism Spectrum Disorders. Autism Res. 9, 951–958 (2016).

18. Liew, Z., Bach, C. C., Asarnow, R. F., Ritz, B. & Olsen, J. Paracetamol use during pregnancy and attention and executive function in offspring at age 5 years. Int. J. Epidemiol. 45, 2009–2017 (2016).

19. Liew, Z. et al. Use of negative control exposure analysis to evaluate confounding: An example of acetaminophen exposure and attention-deficit/hyperactivity disorder in Nurses’ Health Study II. Am. J. Epidemiol. 188, 768–775 (2019).

20. Stergiakouli, E., Thapar, A. & Davey Smith, G. Association of acetaminophen use during pregnancy with behavioral problems in childhood: Evidence against confounding: Evidence against confounding. JAMA Pediatr. 170, 964–970 (2016).

21. Sznajder, K. K., Teti, D. M. & Kjerulff, K. H. Maternal use of acetaminophen during pregnancy and neurobehavioral problems in offspring at 3 years: A prospective cohort study. PLoS One 17, e0272593 (2022).

22. Tovo-Rodrigues, L. et al. Is intrauterine exposure to acetaminophen associated with emotional and hyperactivity problems during childhood? Findings from the 2004 Pelotas birth cohort. BMC Psychiatry 18, 368 (2018).

23. Tovo-Rodrigues, L. et al. Low neurodevelopmental performance and behavioural/emotional problems at 24 and 48 months in Brazilian children exposed to acetaminophen during foetal development. Paediatr. Perinat. Epidemiol. 34, 278–286 (2020).

24. Ystrom, E. et al. Prenatal exposure to acetaminophen and risk of ADHD. Pediatrics 140, e20163840 (2017).

25. Inoue, K. et al. Behavioral problems at age 11 years after prenatal and postnatal exposure to acetaminophen: Parent-reported and self-reported outcomes. Am. J. Epidemiol. 190, 1009–1020 (2021).

26. Ruisch, I. H., Buitelaar, J. K., Glennon, J. C., Hoekstra, P. J. & Dietrich, A. Pregnancy risk factors in relation to oppositional-defiant and conduct disorder symptoms in the Avon Longitudinal Study of Parents and Children. J. Psychiatr. Res. 101, 63–71 (2018).

27. Skovlund, E., Handal, M., Selmer, R., Brandlistuen, R. E. & Skurtveit, S. Language competence and communication skills in 3-year-old children after prenatal exposure to analgesic opioids: Development after Prenatal Opioid Exposure. Pharmacoepidemiol. Drug Saf. 26, 625–634 (2017).

28. Trønnes, J. N., Wood, M., Lupattelli, A., Ystrom, E. & Nordeng, H. Prenatal paracetamol exposure and neurodevelopmental outcomes in preschool-aged children. Paediatr. Perinat. Epidemiol. 34, 247–256 (2020).

29. Vlenterie, R. et al. Neurodevelopmental problems at 18 months among children exposed to paracetamol in utero: a propensity score matched cohort study. Int. J. Epidemiol. 45, 1998–2008 (2016).

30. Avella-Garcia, C. B. et al. Acetaminophen use in pregnancy and neurodevelopment: attention function and autism spectrum symptoms. Int. J. Epidemiol. 45, 1987–1996 (2016).

31. Baker, B. H. et al. Association of prenatal acetaminophen exposure measured in meconium with risk of attention-deficit/hyperactivity disorder mediated by frontoparietal network brain connectivity. JAMA Pediatr. 174, 1073–1081 (2020).

32. Baker, B. H. et al. Associations of maternal blood biomarkers of prenatal APAP exposure with placental gene expression and child attention deficit hyperactivity disorder. Nat. Ment. Health 3, 318–331 (2025).

33. Ji, Y. et al. Association of cord plasma biomarkers of in utero acetaminophen exposure with risk of attention-deficit/hyperactivity disorder and autism spectrum disorder in childhood. JAMA Psychiatry 77, 180–189 (2020).

34. Chen, M.-H. et al. Prenatal exposure to acetaminophen and the risk of attention-deficit/hyperactivity disorder: A nationwide study in Taiwan: A nationwide study in Taiwan. J. Clin. Psychiatry 80, 18m12612 (2019).

35. Bornehag, C.-G. et al. Prenatal exposure to acetaminophen and children’s language development at 30 months. Eur. Psychiatry 51, 98–103 (2018).

36. Mueller, K. F. et al. Methods for detecting, quantifying, and adjusting for dissemination bias in meta-analysis are described. J. Clin. Epidemiol. 80, 25–33 (2016).

37. Marks-Anglin, A. & Chen, Y. A historical review of publication bias. Res. Synth. Methods 11, 725–742 (2020).

38. Duval, S. & Tweedie, R. Trim and fill: A simple funnel-plot-based method of testing and adjusting for publication bias in meta-analysis. Biometrics 56, 455–463 (2000).

39. Duval, S. & Tweedie, R. A nonparametric ‘trim and fill’ method of accounting for publication bias in meta-analysis. J. Am. Stat. Assoc. 95, 89–98 (2000).

40. Simonsohn, U., Nelson, L. D. & Simmons, J. P. P-curve: a key to the file-drawer. J. Exp. Psychol. Gen. 143, 534–547 (2014).

41. Hedges, L. V. Modeling publication selection effects in meta-analysis. JSTOR 7, 246–255 (1992).

42. Iyengar, S. & Greenhouse, J. B. Selection models and the file drawer problem. Stat. Sci. 3, 109–117 (1988).

43. Vevea, J. L. & Hedges, L. V. A general linear model for estimating effect size in the presence of publication bias. Psychometrika 60, 419–435 (1995).

44. Rücker, G., Schwarzer, G., Carpenter, J. R., Binder, H. & Schumacher, M. Treatment-effect estimates adjusted for small-study effects via a limit meta-analysis. Biostatistics 12, 122–142 (2011).

45. Dear, K. B. G. & Begg, C. B. An approach for assessing publication bias prior to performing a meta-analysis. Stat. Sci. 7, 237–245 (1992).

46. Copas, J. B. & Shi, J. Q. A sensitivity analysis for publication bias in systematic reviews. Stat. Methods Med. Res. 10, 251–265 (2001).

47. van Assen, M. A. L. M., van Aert, R. C. M. & Wicherts, J. M. Meta-analysis using effect size distributions of only statistically significant studies. Psychol. Methods 20, 293–309 (2015).

48. Mathur, M. B. & VanderWeele, T. J. Sensitivity analysis for unmeasured confounding in meta-analyses. J. Am. Stat. Assoc. 115, 163–172 (2020).

49. Salanti, G. & Ioannidis, J. P. A. Synthesis of observational studies should consider credibility ceilings. J. Clin. Epidemiol. 62, 115–122 (2009).

50. Danielson, M. L. et al. ADHD prevalence among U.s. children and adolescents in 2022: Diagnosis, severity, co-occurring disorders, and treatment. J. Clin. Child Adolesc. Psychol. 53, 343–360 (2024).

51. Maenner, M. J. et al. Prevalence and characteristics of Autism spectrum disorder among children aged 8 years - Autism and Developmental Disabilities Monitoring network, 11 sites, United States, 2020. MMWR Surveill. Summ. 72, 1–14 (2023).

52. VanderWeele, T. J. Optimal approximate conversions of odds ratios and hazard ratios to risk ratios. Biometrics 76, 746–752 (2020).

53. Luo, S. et al. Prenatal acetaminophen (paracetamol) use and the risk of autism and/or attention-deficit/hyperactivity disorder among sibling-matched cohorts. JAMA Intern. Med. (2026) doi:10.1001/jamainternmed.2026.2215.

54. Brandlistuen, R. E., Ystrom, E., Nulman, I., Koren, G. & Nordeng, H. Prenatal paracetamol exposure and child neurodevelopment: a sibling-controlled cohort study. Int. J. Epidemiol. 42, 1702–1713 (2013).

55. Masarwa, R. et al. Prenatal exposure to acetaminophen and risk for attention deficit hyperactivity disorder and autistic spectrum disorder: A systematic review, meta-analysis, and meta-regression analysis of cohort studies. Am. J. Epidemiol. 187, 1817–1827 (2018).

56. Ricci, C. et al. In utero acetaminophen exposure and child neurodevelopmental outcomes: Systematic review and meta-analysis. Paediatr. Perinat. Epidemiol. 37, 473–484 (2023).

57. D’Antonio, F. et al. Prenatal paracetamol exposure and child neurodevelopment: a systematic review and meta-analysis. Lancet Obstet. Gynaecol. Womens Health 2, e190–e198 (2026).

58. Bérard, A. et al. Systematic Review and meta-analysis: Acetaminophen use during pregnancy and the risk of neurodevelopmental disorders in childhood. J. Am. Acad. Child Adolesc. Psychiatry 65, 484–504 (2026).

59. Boccia, S. et al. Registration practices for observational studies on ClinicalTrials.gov indicated low adherence. J. Clin. Epidemiol. 70, 176–182 (2016).

60. Janiaud, P. et al. Validity of observational evidence on putative risk and protective factors: appraisal of 3744 meta-analyses on 57 topics. BMC Med. 19, 157 (2021).

61. Yaacoub, S. et al. Factors influencing the trustworthiness of non-randomized studies of interventions: a survey of international experts. BMC Med. (2026) doi:10.1186/s12916-026-04979-6.

62. Bruns, S. B. & Ioannidis, J. P. A. P-curve and p-hacking in observational research. PLoS One 11, e0149144 (2016).

